# KESOZI Digital Twin: Physics-Informed Neural Network for Independent Estimation and Prediction of Childhood Diarrheal Disease Burden in Kenya, Somaliland, and Zimbabwe

**DOI:** 10.64898/2026.06.03.26354823

**Authors:** John Onyango Agumba, Lucy Namusonge, Joshua Ogendo, Musiiwa Takavarasha, Mohamad Ahmed Hassan, Morris Senghor, Lydia Waswa, Anthony Pembere

**Author notes:** Corresponding Author: John Agumba.

## Abstract

Childhood diarrheal disease remains a leading cause of morbidity and mortality among children under five years in sub-Saharan Africa, particularly in settings affected by inadequate sanitation, climate variability, malnutrition, and limited healthcare access. Conventional forecasting approaches are often constrained by sparse surveillance data, weak spatial representation, and limited incorporation of mechanistic disease dynamics. This study presents a Physics-Informed Multimodal Artificial Intelligence Digital Twin framework that integrates Physics-Informed Neural Networks, Graph Neural Networks, diffusion-reaction epidemiological modeling, multimodal fusion learning, and Digital Twin simulation to estimate and predict childhood diarrheal disease burden in Kenya, Somaliland, and Zimbabwe. Using public epidemiological, environmental, climate, sanitation, and synthetic proof-of-concept datasets, the framework modeled temporal disease dynamics, spatial transmission, pathogen-attributed burden, and outbreak trajectories while enforcing epidemiological consistency through physics-informed optimization. Results demonstrated robust forecasting performance, enhanced spatial transmission modeling, uncertainty-aware predictions, and realistic outbreak simulations across the three countries. Rotavirus, Shigella, and Cryptosporidium were identified as major contributors to modeled mortality burden, while unsafe water exposure, poor sanitation, malnutrition, and climate-sensitive transmission substantially increased disease risk. Compared with a Bayesian baseline model, the multimodal framework achieved superior nonlinear risk characterization, geospatial learning, and temporal prediction. These findings highlight the potential of scientific machine learning and digital twin systems for infectious disease surveillance, outbreak forecasting, climate-health analytics, and evidence-based public health decision-making in low-resource African settings.

## 1. Introduction

Childhood diarrheal disease remains among the leading causes of mortality and morbidity among children under five years globally despite substantial improvements in sanitation, vaccination, nutrition, and healthcare interventions. The burden remains disproportionately high in sub-Saharan Africa, where climate-sensitive transmission, poor sanitation infrastructure, limited healthcare access, malnutrition, and unsafe drinking water continue to amplify disease risk. Kenya, Somaliland, and Zimbabwe continue to experience substantial spatial and environmental heterogeneity in diarrheal disease transmission, with significant variability in disease burden between urban, rural, arid, flood-prone, and underserved regions. According to the World Health Organization, diarrheal diseases remain among the leading causes of childhood mortality in low-resource settings, particularly in regions characterized by weak sanitation systems and limited healthcare infrastructure (World Health Organization [WHO], 2024). The epidemiology of diarrheal disease involves complex interactions among environmental conditions, sanitation systems, climate variability, healthcare accessibility, nutrition, population density, pathogen transmission dynamics, and regional mobility patterns. These nonlinear interactions create substantial challenges for conventional epidemiological forecasting systems, which often struggle to capture spatial transmission dependencies, temporal outbreak dynamics, and environmental-health interactions. Furthermore, many existing epidemiological machine-learning systems remain primarily data-driven and fail to incorporate governing epidemiological dynamics and physical propagation laws, thereby limiting interpretability and reducing epidemiological consistency under sparse or noisy surveillance conditions commonly observed in low-resource healthcare environments.

Artificial intelligence has increasingly emerged as an important tool for infectious disease surveillance, epidemiological forecasting, and healthcare analytics. Machine-learning approaches including deep neural networks, recurrent neural networks, transformers, and ensemble learning methods have demonstrated promising performance in predicting disease outbreaks, transmission dynamics, and healthcare risk patterns (Esteva et al., 2019). AI-driven epidemiological systems have been successfully applied in influenza prediction, COVID-19 outbreak forecasting, tuberculosis screening, and climate-sensitive disease modeling. However, many existing approaches remain weakly integrated with mechanistic epidemiological dynamics and physical disease-propagation laws. Consequently, such systems may generate physically inconsistent predictions, particularly when operating under incomplete surveillance conditions or rapidly evolving outbreak environments. Recent advances in scientific machine learning have enabled development of hybrid frameworks capable of integrating physical laws with artificial intelligence systems. Physics-Informed Neural Networks (PINNs) incorporate governing differential equations directly into neural network optimization, thereby improving physical consistency, stability, and generalization (Raissi et al., 2019). PINNs have demonstrated effectiveness in scientific systems characterized by sparse, noisy, or incomplete observations because the learning process is constrained by governing physical equations rather than relying exclusively on observational data. Their ability to integrate mechanistic disease dynamics into machine-learning optimization makes them particularly suitable for epidemiological systems involving nonlinear transmission behavior, climate-sensitive propagation, and incomplete surveillance reporting.

In parallel, Graph Neural Networks (GNNs) have emerged as important tools for spatial epidemiological modeling. GNN architectures are specifically designed for graph-structured systems in which nodes and edges represent interacting entities and transmission relationships. In epidemiological applications, nodes may represent counties, healthcare regions, or communities, while edges may represent transportation systems, geographical proximity, environmental similarity, or mobility-driven interactions (Kapoor et al., 2020). Graph-based learning frameworks therefore enable modeling of spatial clustering behavior, neighboring transmission influence, and regional outbreak propagation. Nevertheless, many current graph-based epidemiological systems remain weakly integrated with governing epidemiological equations and dynamic outbreak-simulation frameworks. Digital Twin systems have additionally emerged as powerful frameworks for real-time monitoring, dynamic simulation, predictive analysis, and intervention-response modeling. Originally developed within industrial and engineering systems, digital twins are increasingly being explored in healthcare and epidemiology for outbreak simulation, healthcare-resource optimization, and real-time disease forecasting (Fuller et al., 2020). Epidemiological digital twins provide opportunities for continuously integrating surveillance data, environmental indicators, climate variables, and predictive artificial intelligence models into dynamic simulation environments capable of modeling evolving epidemiological conditions. Despite their growing potential, digital twin applications remain limited in low-resource healthcare environments, particularly for childhood infectious disease surveillance in Africa.

Despite increasing adoption of artificial intelligence in healthcare analytics, limited work has integrated epidemiological physics, graph learning, multimodal fusion modeling, pathogen-attribution analytics, uncertainty-aware forecasting, and digital twin systems for childhood diarrheal disease surveillance in Africa. Existing epidemiological forecasting systems are frequently constrained by sparse surveillance data, weak spatial modeling capability, limited physical interpretability, poor uncertainty characterization, and inadequate real-time simulation functionality. Many current models rely heavily on purely data-driven learning approaches that fail to preserve epidemiological consistency during forecasting and do not adequately capture regional transmission interactions, pathogen-attributed burden, environmental-health coupling, or dynamically evolving outbreak conditions.

To address these limitations, this study develops a Physics-Informed Multimodal Artificial Intelligence Digital Twin framework integrating Physics-Informed Neural Networks, diffusion-reaction epidemiological equations, Graph Neural Networks, multimodal fusion learning, pathogen-attribution modeling, geospatial epidemiology, uncertainty calibration, and Digital Twin simulation for independent estimation and prediction of childhood diarrheal disease burden in Kenya, Somaliland, and Zimbabwe. The proposed framework integrates epidemiological, climate, environmental, sanitation, healthcare-access, nutritional, and pathogen-related datasets to model temporal disease dynamics, spatial propagation behavior, outbreak evolution, and intervention-response scenarios. By combining scientific machine learning with multimodal epidemiological fusion and digital twin simulation, the framework aims to improve disease-burden estimation, forecasting stability, hotspot identification, pathogen-attributed burden analysis, and evidence-based public-health decision support in resource-constrained African settings.

## 2. Materials and Methods

### 2.1 Overall Framework Architecture

The proposed framework integrates epidemiological diffusion-reaction modeling, Physics-Informed Neural Networks, Graph Neural Networks, multimodal epidemiological datasets, pathogen attribution analytics, uncertainty calibration, and Digital Twin simulation architecture to model childhood diarrheal disease burden and transmission dynamics in Kenya, Somaliland, and Zimbabwe.

### 2.2 Data Sources and Preprocessing

Publicly available datasets were obtained from the World Health Organization, World Bank, UNICEF, and Our World in Data. The integrated datasets included under-five mortality rates, diarrheal disease indicators, climate variables, healthcare accessibility indicators, water and sanitation indicators, and population statistics relevant to childhood diarrheal disease transmission and burden estimation.

Synthetic proof-of-concept datasets additionally incorporated under-five population estimates, diarrheal cases, severe disease cases, hospitalizations, mortality data, climate variables, WASH indicators, nutrition-risk indicators, healthcare-access indicators, pathogen prevalence estimates, satellite-derived environmental-risk scores, spatial coordinates, and temporal seasonal variables to support multimodal epidemiological modeling, spatial transmission analysis, pathogen-attribution estimation, climate-health interaction modeling, and digital twin outbreak simulation across Kenya, Somaliland, and Zimbabwe. The modeled pathogens included Rotavirus, Shigella, Adenovirus 40/41, Norovirus GII, ST-ETEC, Cholera, Cryptosporidium, and Campylobacter.

### 2.3 Epidemiological and Physical Modeling Framework

#### 2.3.1 Diffusion-Reaction Epidemiological Equation

The governing epidemiological dynamics were modeled using:

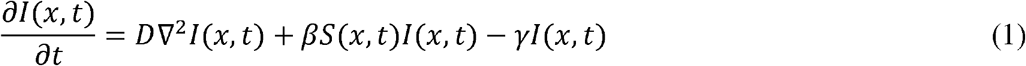

where *I* (*x,t*) represents infected individuals at spatial position *x* and time *t, S*(*x,t*) represents, susceptible individuals, *D* denotes the spatial diffusion coefficient, *β* represents disease transmission rate and *γ* represents recovery rate.

#### 2.3.2 Spatial Diffusion Equation

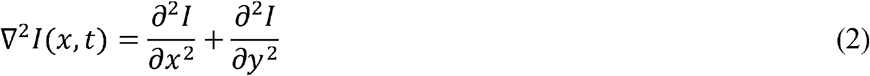

#### 2.3.3 Susceptible-Infected--Recovered Dynamics

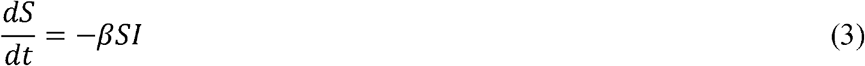

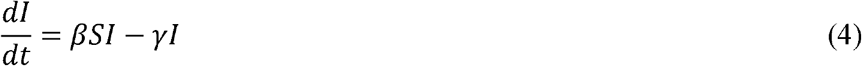

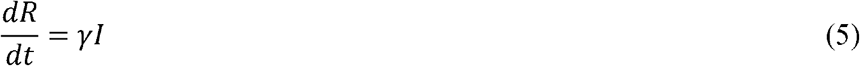

#### 2.3.4 Disease Burden Estimation Equation

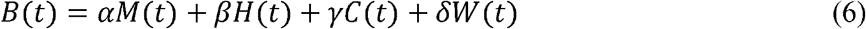

where *B*(*t*) represents total disease burden, *M*(*t*) represents mortality indicators, *H*(*t*) represents healthcare indicators, *C*(*t*) represents climate indicators and *W*(*t*) represents WASH indicators

### 2.4 Physics-Informed Neural Network Framework

The total PINN optimization loss was represented as:

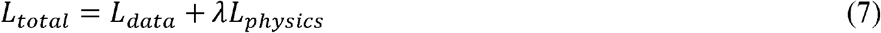

The observational data loss was expressed as:

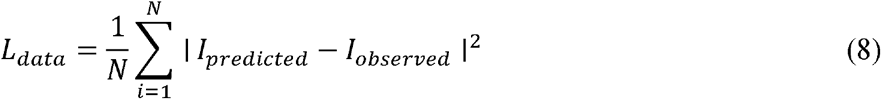

The epidemiological physics loss was defined as:

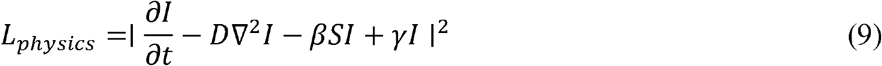

### 2.5 Multimodal Fusion Framework

The multimodal fusion framework combined Bayesian baseline modeling, boosting models, temporal models, geospatial models, satellite environmental-risk models, and pathogen-attribution analytics.

The fusion model was represented as:

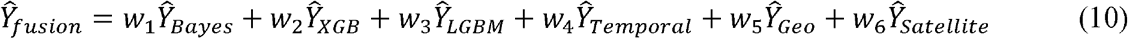

subject to:

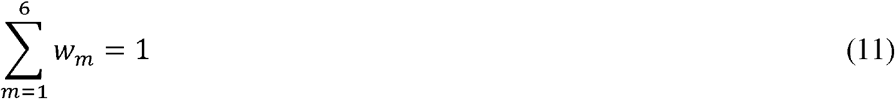

### 2.6 Graph Neural Network Spatial Framework

The graph aggregation mechanism was represented as:

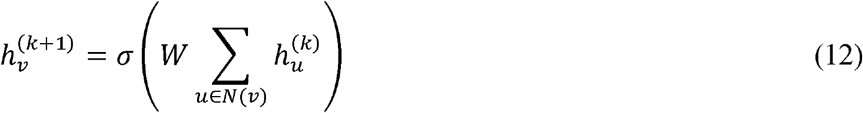

Where 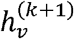 represents updated embedding of region 𝒱, *N*(𝒱)denotes neighboring regions connected to region 𝒱 and *W* represents trainable graph weights.

## 3. Results

### 3.1 Synthetic Multimodal Epidemiological Data Generation and Cross-Country Disease Dynamics

Figure 2 shows the relationship between the composite diarrheal disease risk score and under-five diarrheal incidence across Kenya, Somaliland, and Zimbabwe. A strong positive association was observed, with higher risk scores corresponding to increased diarrheal incidence rates. Somaliland generally exhibited higher risk scores and incidence levels than the other countries, while Kenya and Zimbabwe showed broader distributions across moderate-risk conditions. The clustering pattern indicates that locations with elevated environmental, nutritional, healthcare, and WASH-related risks experienced substantially higher diarrheal disease incidence.

**Figure 2.**
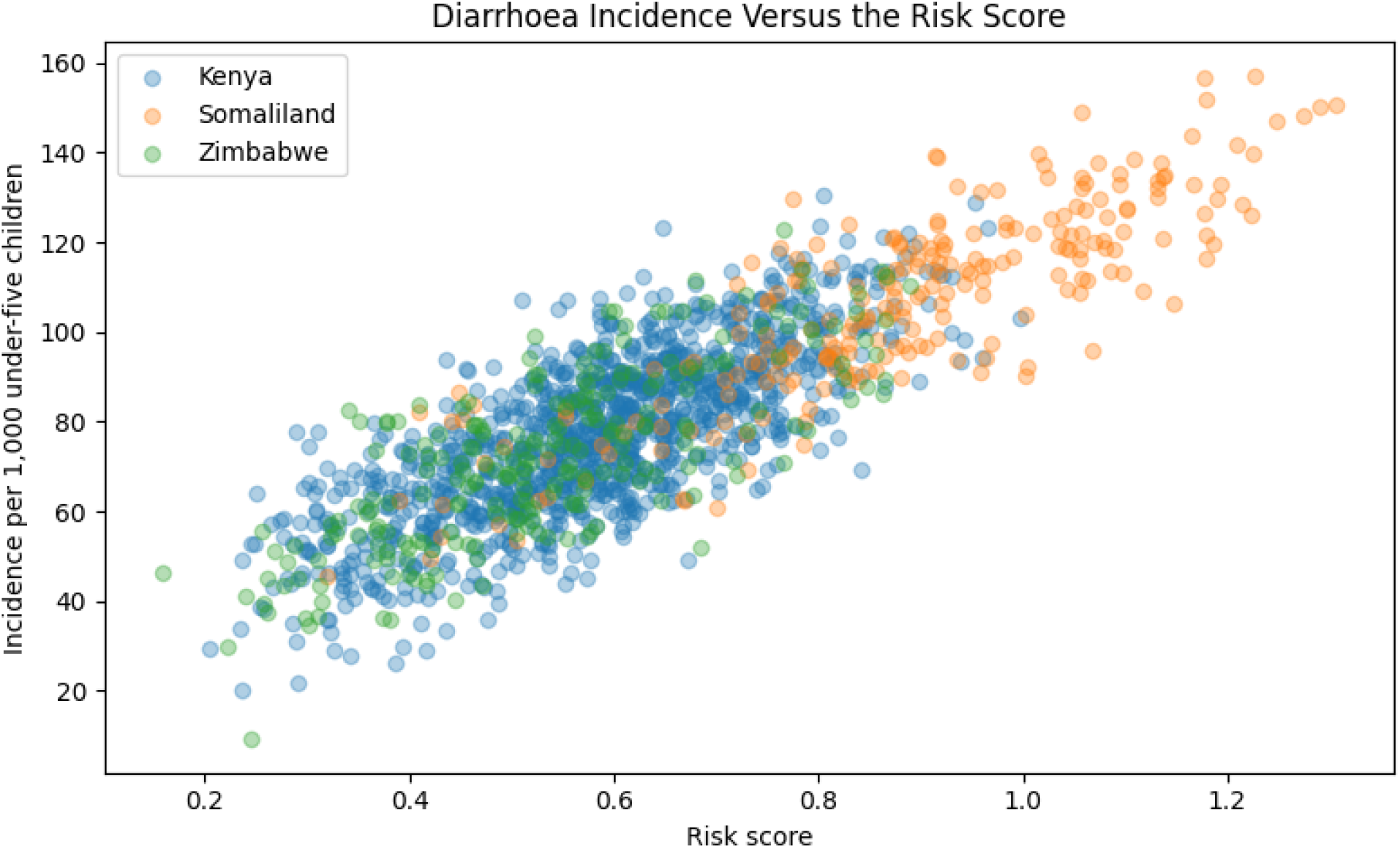
Relationship between composite risk score and under-five diarrheal incidence across Kenya, Somaliland, and Zimbabwe. Higher risk scores were associated with increased diarrheal incidence, demonstrating the influence of combined environmental, healthcare, nutritional, and WASH-related risk factors on disease burden.

### 3.2. Climate Forcing Dynamics Across Study Settings

Figure 3 compares the modeled climate forcing across Kisumu (Kenya), Hargeisa (Somaliland), and Harare (Zimbabwe) between January 2024 and December 2025. Substantial temporal variability was observed in all three locations, reflecting fluctuations in climatic conditions that influence diarrheal disease transmission. Kisumu exhibited the highest peak climate forcing during mid-2025, while Harare showed several elevated forcing periods throughout the study period. Hargeisa demonstrated moderate but persistent variability, with periodic increases in climate forcing. These patterns indicate that climatic influences on diarrheal disease risk vary considerably across both time and geographic location.

**Figure 3.**
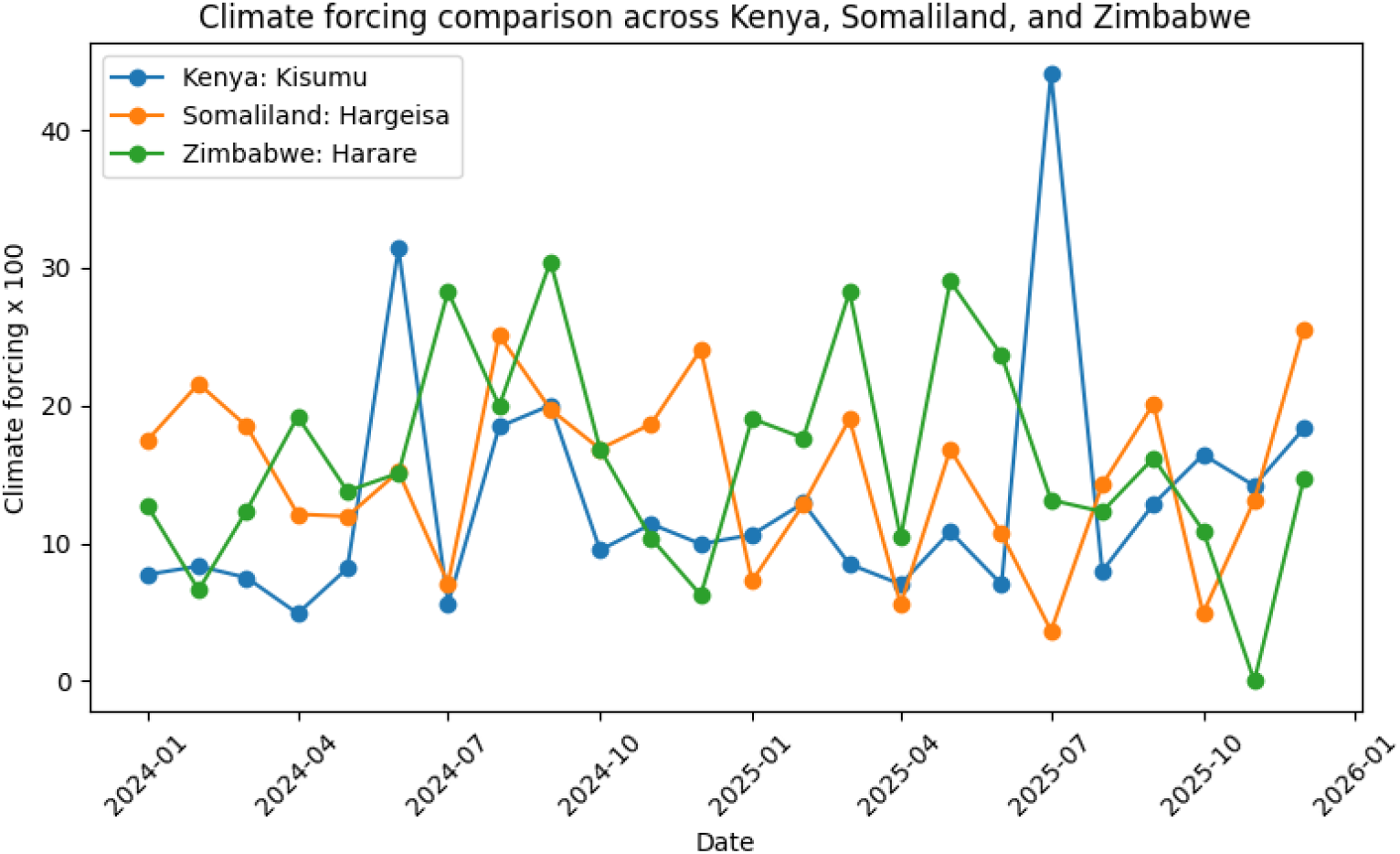
Climate forcing trends across Kenya, Somaliland, and Zimbabwe (2024-2025). Monthly climate forcing values for Kisumu, Hargeisa, and Harare showing temporal variability in climatic conditions that may influence diarrheal disease transmission and burden.

### 3.3 Country-Level Diarrheal Mortality Burden

Figure 4 presents the estimated under-five diarrheal mortality burden across the study countries in 2025. Kenya exhibited the highest modeled mortality burden, with approximately 2,400 diarrheal deaths, followed by Somaliland with approximately 700 deaths and Zimbabwe with approximately 400 deaths. The results indicate substantial variation in mortality burden across the three settings, reflecting differences in population size, environmental risk factors, healthcare access, sanitation conditions, and other determinants incorporated within the KESOZI Digital Twin framework.

**Figure 4.**
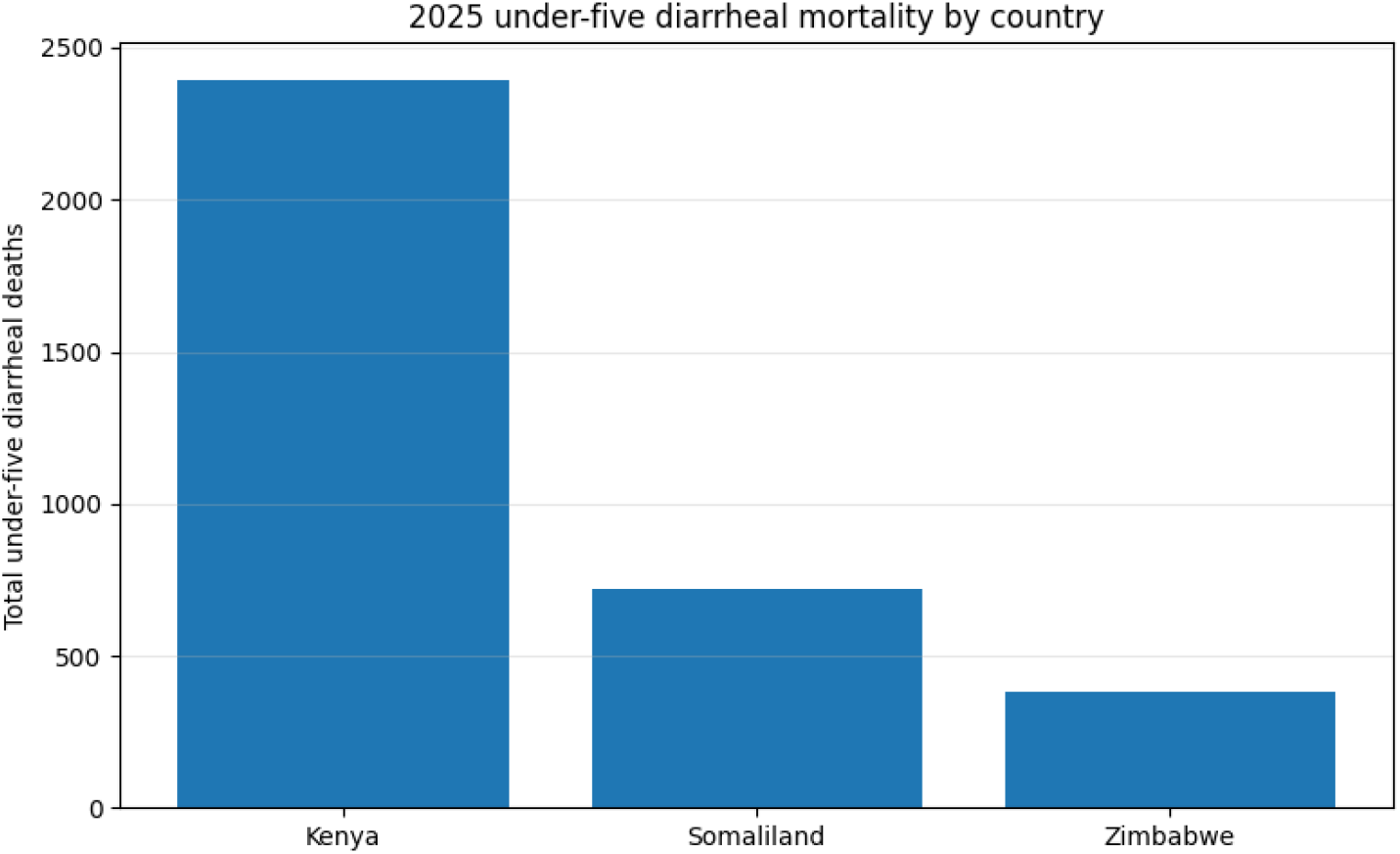
Estimated under-five diarrheal deaths by country in 2025. Country-level modeled diarrheal mortality burden among children under five years of age generated using the KESOZI Digital Twin framework.

### 3.4 Mortality Incidence Rates Across Study Countries

Figure 5 presents the estimated under-five diarrheal mortality incidence rates across Kenya, Somaliland, and Zimbabwe in 2025. Somaliland exhibited the highest mortality incidence rate, at approximately 4.1 deaths per 100,000 under-five children, followed by Kenya at approximately 1.9 deaths per 100,000 and Zimbabwe at approximately 1.6 deaths per 100,000. Although Kenya recorded the highest absolute number of diarrheal deaths, Somaliland experienced the greatest mortality burden relative to its under-five population, indicating a higher risk of diarrheal death among children under five.

**Figure 5.**
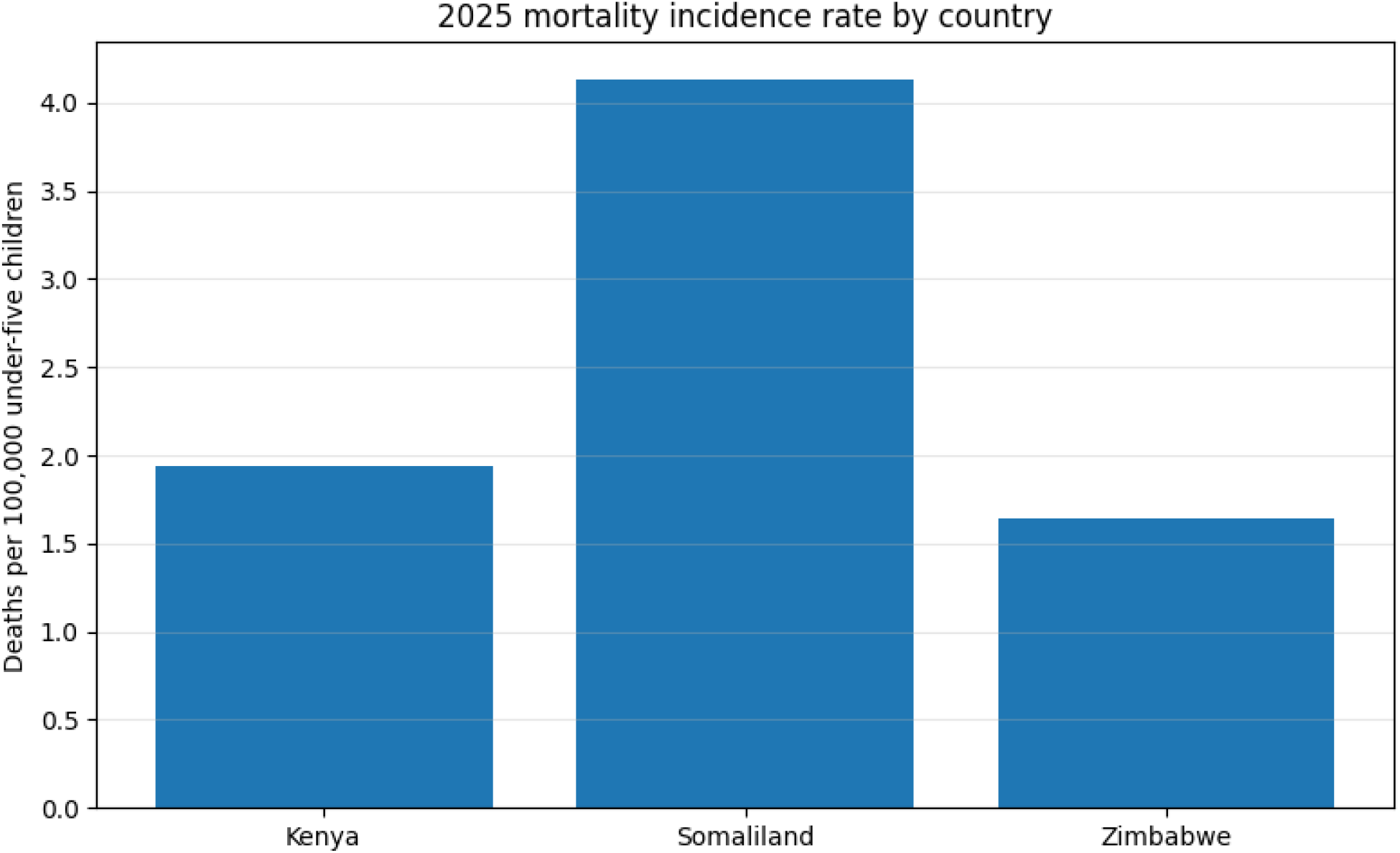
Under-five diarrheal mortality incidence rates by country in 2025. Estimated diarrheal deaths per 100,000 under-five children in Kenya, Somaliland, and Zimbabwe generated using the KESOZI Digital Twin framework.

### 3.5 Morbidity Incidence Rates Across the Study Countries

Figure 6 presents the estimated under-five diarrheal morbidity incidence rates across Kenya, Somaliland, and Zimbabwe in 2025. Somaliland exhibited the highest morbidity incidence rate, with approximately 9.3 cases per 1,000 under-five children, followed by Kenya at approximately 6.4 cases per 1,000 and Zimbabwe at approximately 5.7 cases per 1,000. These results indicate substantial variation in diarrheal disease occurrence across the study settings, with children in Somaliland experiencing the highest population-adjusted burden of diarrheal illness.

**Figure 6.**
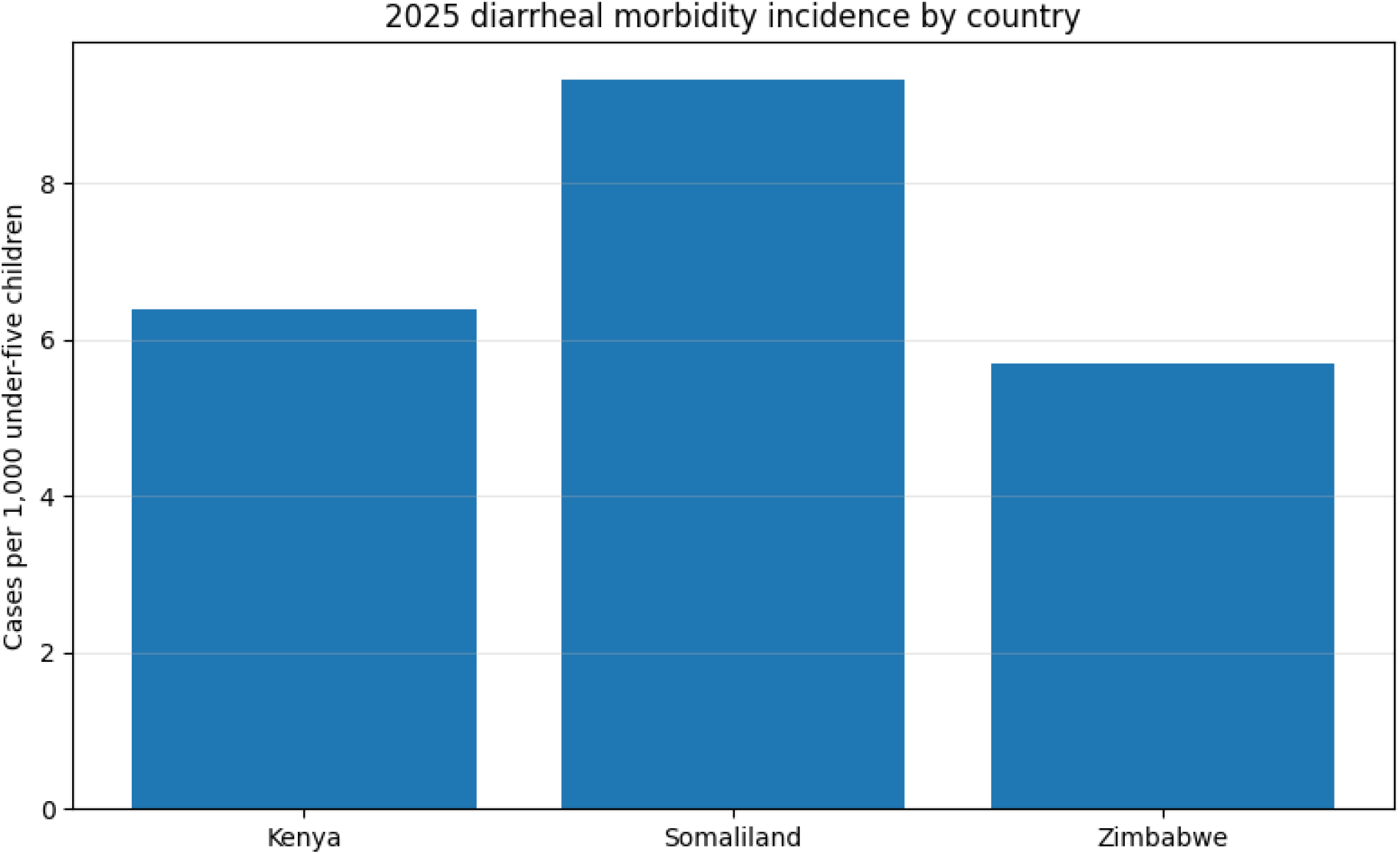
Under-five diarrheal morbidity incidence rates by country in 2025. Estimated diarrheal cases per 1,000 under-five children in Kenya, Somaliland, and Zimbabwe generated using the KESOZI Digital Twin framework.

### 3.6 Comparative Diarrheal Burden Components Across Study Countries

Figure 7 compares the major components of the modeled under-five diarrheal disease burden across Kenya, Somaliland, and Zimbabwe in 2025, including total cases, severe cases, hospitalizations, and deaths. Kenya exhibited the highest overall burden across all indicators, recording the greatest number of diarrheal cases, severe cases, hospitalizations, and deaths. Somaliland showed an intermediate burden, while Zimbabwe consistently recorded the lowest counts across the burden components. The results demonstrate substantial differences in disease occurrence, severity, healthcare utilization, and mortality among the three study settings.

**Figure 7.**
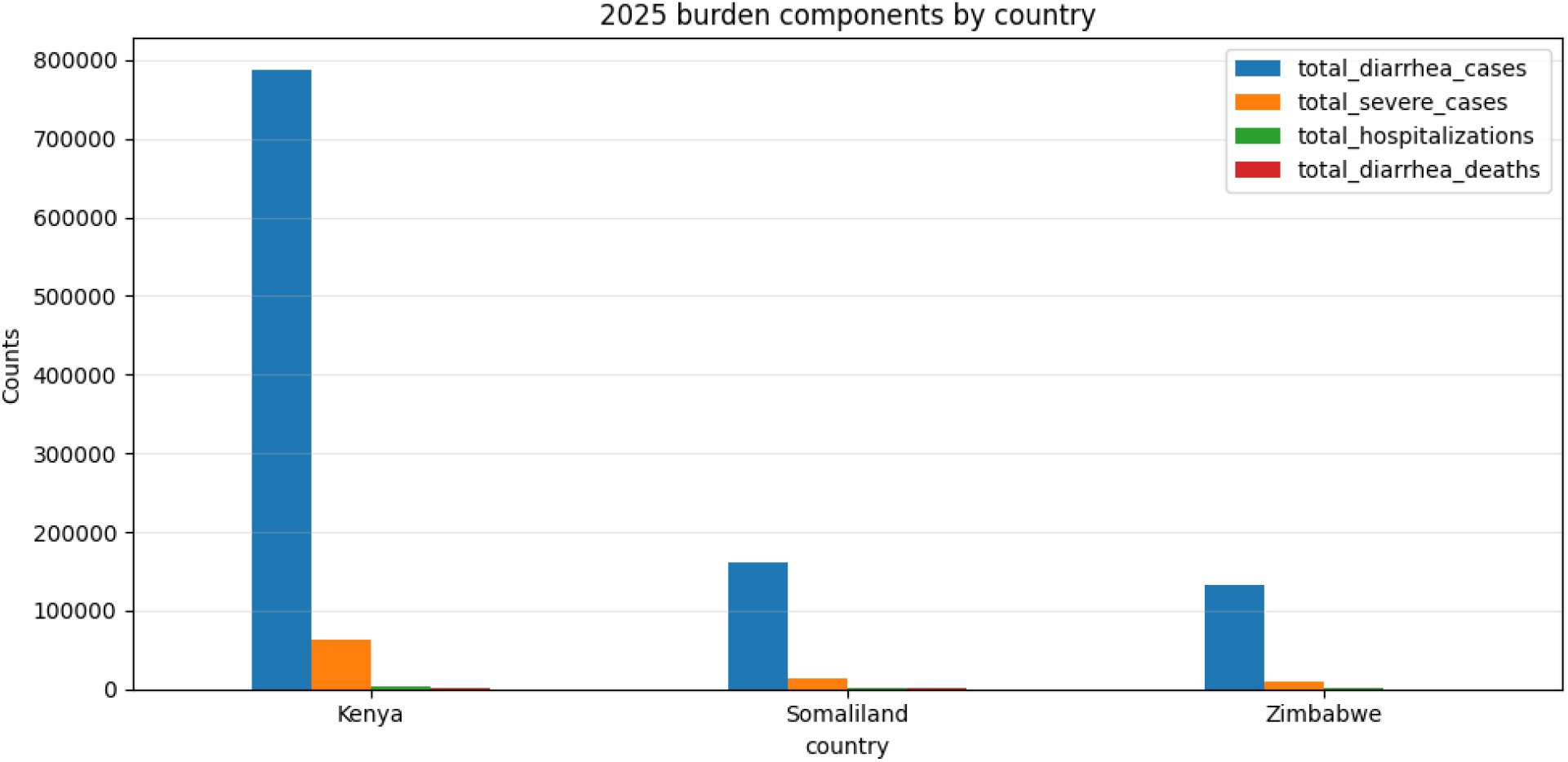
Comparative diarrheal burden components by country in 2025. Estimated total diarrheal cases, severe cases, hospitalizations, and deaths among children under five in Kenya, Somaliland, and Zimbabwe generated using the KESOZI Digital Twin framework.

### 3.7 Comparison of Diarrheal Disease Risk Drivers Across Study Countries

Figure 8 compares the major risk drivers influencing under-five diarrheal disease burden across Kenya, Somaliland, and Zimbabwe. Somaliland exhibited the highest overall risk score, climate forcing, unsafe water exposure, and wasting prevalence, indicating a greater concentration of environmental and nutritional vulnerabilities. Kenya showed intermediate levels for most risk indicators, while Zimbabwe generally exhibited the lowest risk scores and unsafe water exposure. In contrast, Zimbabwe recorded the highest health-access index, followed by Kenya, whereas Somaliland demonstrated the lowest healthcare access among the three settings.

**Figure 8.**
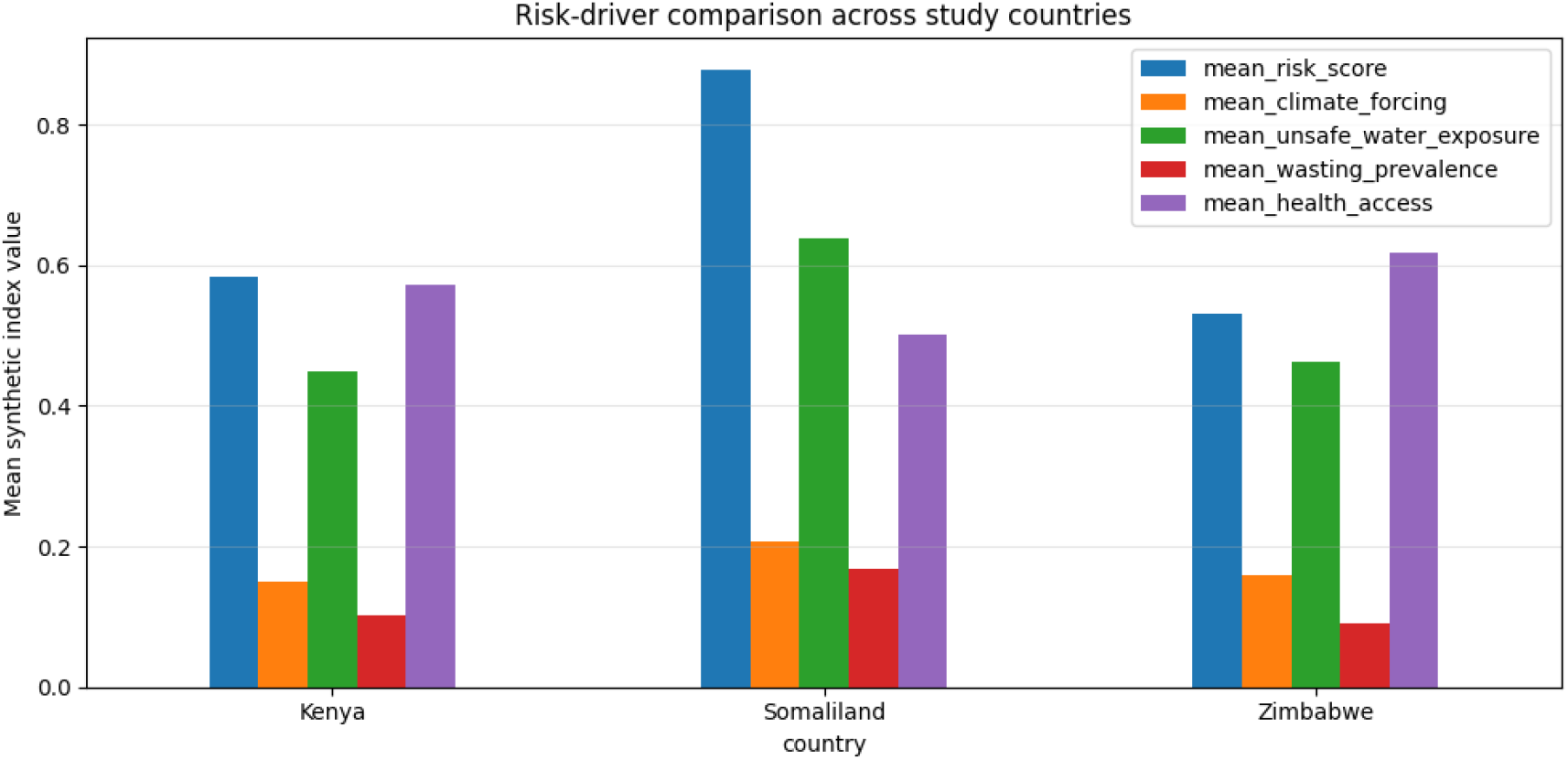
Comparison of major diarrheal disease risk drivers across Kenya, Somaliland, and Zimbabwe. Mean values of composite risk score, climate forcing, unsafe water exposure, wasting prevalence, and healthcare access indicators used in the KESOZI Digital Twin framework.

### 3.8 Temporal Trends in Diarrheal Disease Burden

Figure 9 illustrates the monthly distribution of under-five diarrheal cases across Kenya, Somaliland, and Zimbabwe in 2025. Kenya consistently recorded the highest number of diarrheal cases throughout the year, with incidence increasing from January and peaking between June and August before declining toward the end of the year. Somaliland exhibited moderate seasonal variation, with cases rising gradually during the first half of the year and remaining relatively stable before decreasing slightly during the final months. Zimbabwe recorded the lowest case counts overall but displayed a similar seasonal pattern, with a mid-year peak followed by a gradual decline. All three countries demonstrated clear temporal variability, indicating the influence of seasonal and environmental factors on disease transmission.

**Figure 9.**
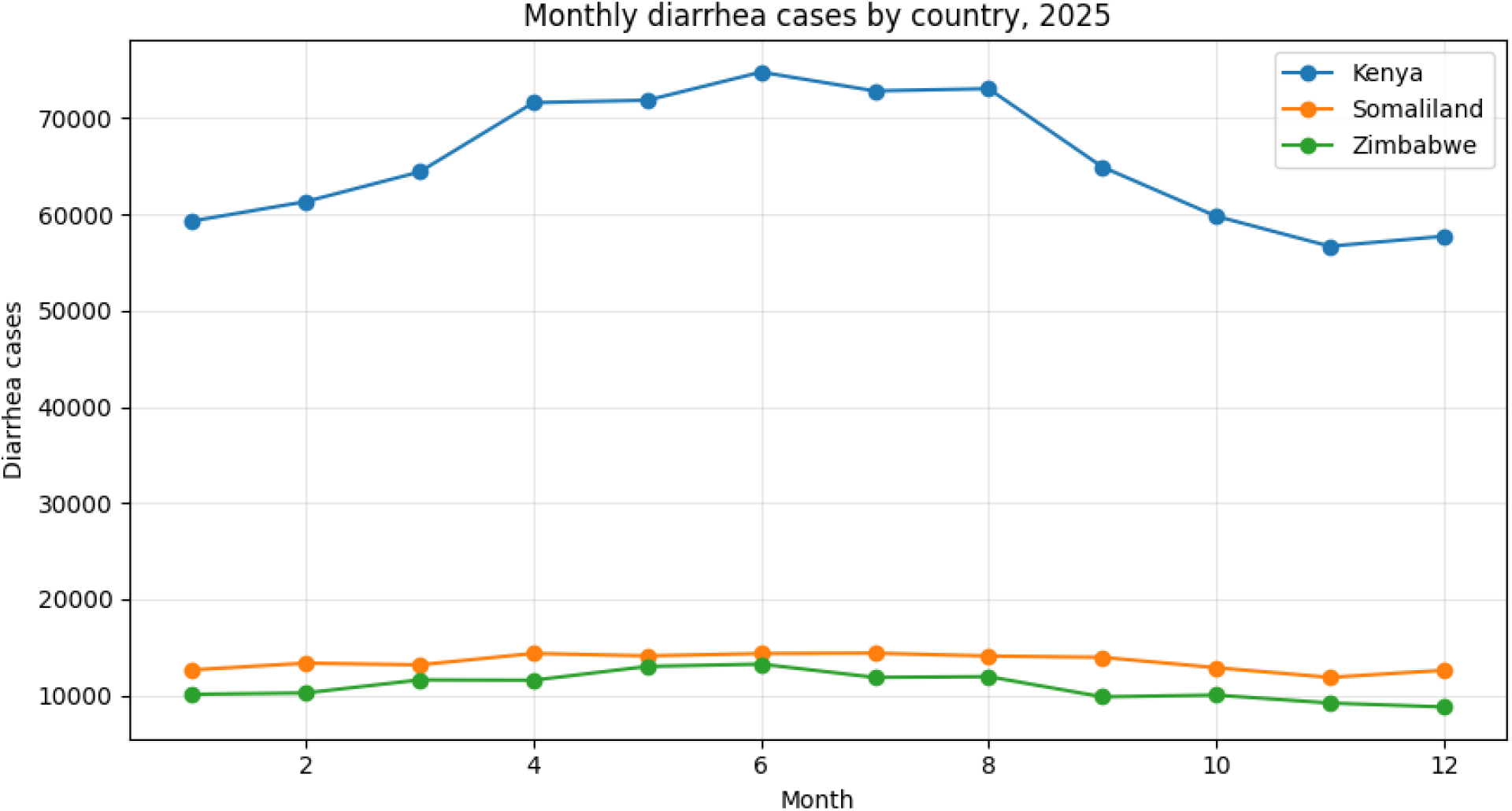
Monthly under-five diarrheal cases across Kenya, Somaliland, and Zimbabwe in 2025. Temporal trends in modeled diarrheal case counts showing seasonal variation and mid-year peaks in disease burden across the study countries.

### 3.9 Pathogen-Specific Mortality Burden by Country

Figure 10 presents pathogen-attributed diarrheal deaths among children under five across Kenya, Somaliland, and Zimbabwe in 2025. Rotavirus accounted for the highest pathogen-attributed mortality burden in all three countries, followed by Shigella and Cryptosporidium. Norovirus GII and ST-ETEC contributed moderate mortality burdens, whereas Cholera, Campylobacter, and Adenovirus 40/41 accounted for comparatively smaller numbers of attributed deaths. Kenya consistently recorded the highest pathogen-attributed mortality across all pathogens, while Somaliland and Zimbabwe exhibited lower but similar mortality profiles. Despite differences in magnitude, the relative ranking of major pathogens remained broadly consistent across the three countries.

**Figure 10.**
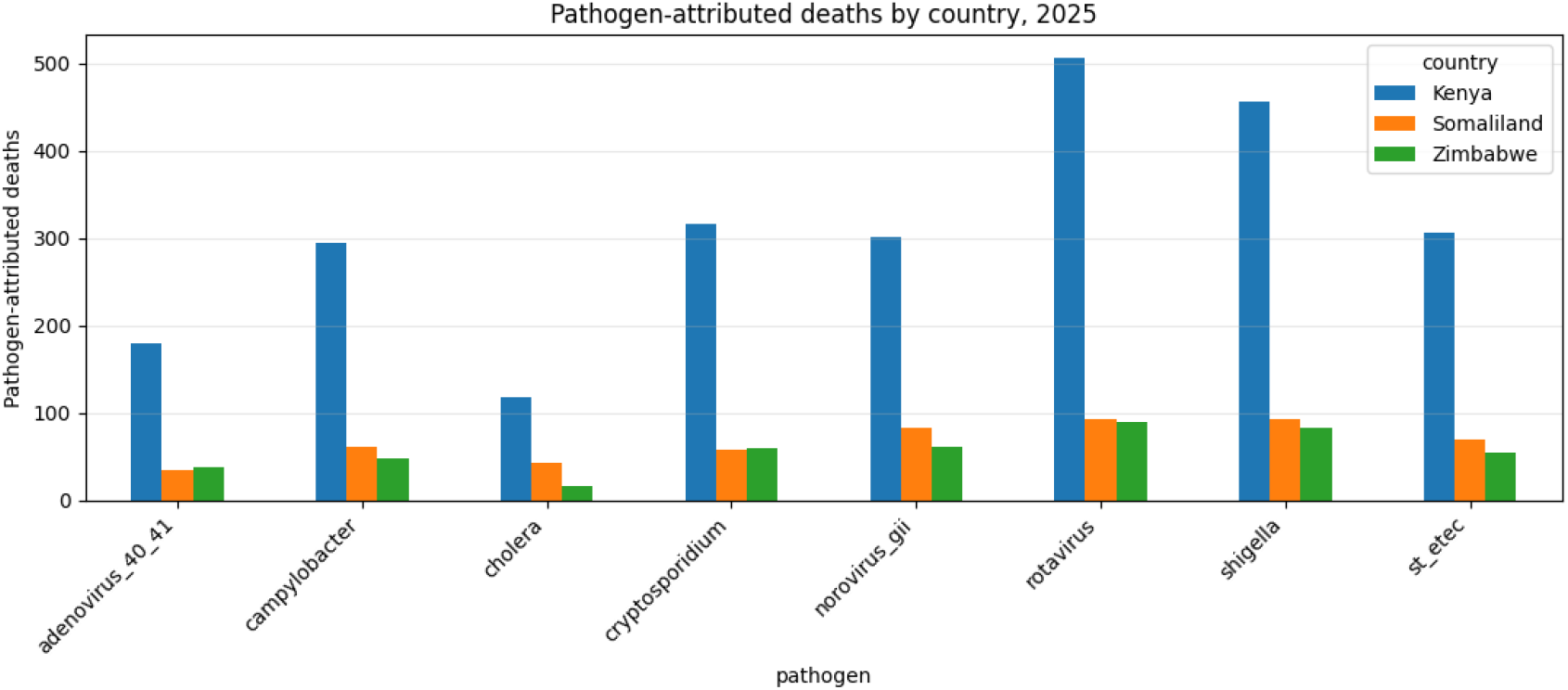
Estimated under-five diarrheal deaths attributed to major enteric pathogens across Kenya, Somaliland, and Zimbabwe generated using the KESOZI Digital Twin framework.

### 3.10 Pathogen-Specific Mortality Fractions

Figure 11 presents the pathogen-specific mortality fractions across Kenya, Somaliland, and Zimbabwe in 2025. Rotavirus accounted for the largest proportion of diarrheal mortality in all three countries, contributing approximately 17-20% of pathogen-attributed deaths, followed by Shigella (17-19%). Cryptosporidium, Norovirus GII, ST-ETEC, and Campylobacter contributed intermediate mortality fractions, while Cholera and Adenovirus 40/41 generally accounted for smaller proportions of mortality. Although the magnitude of individual pathogen contributions varied across countries, the overall mortality attribution patterns were broadly consistent.

**Figure 11.**
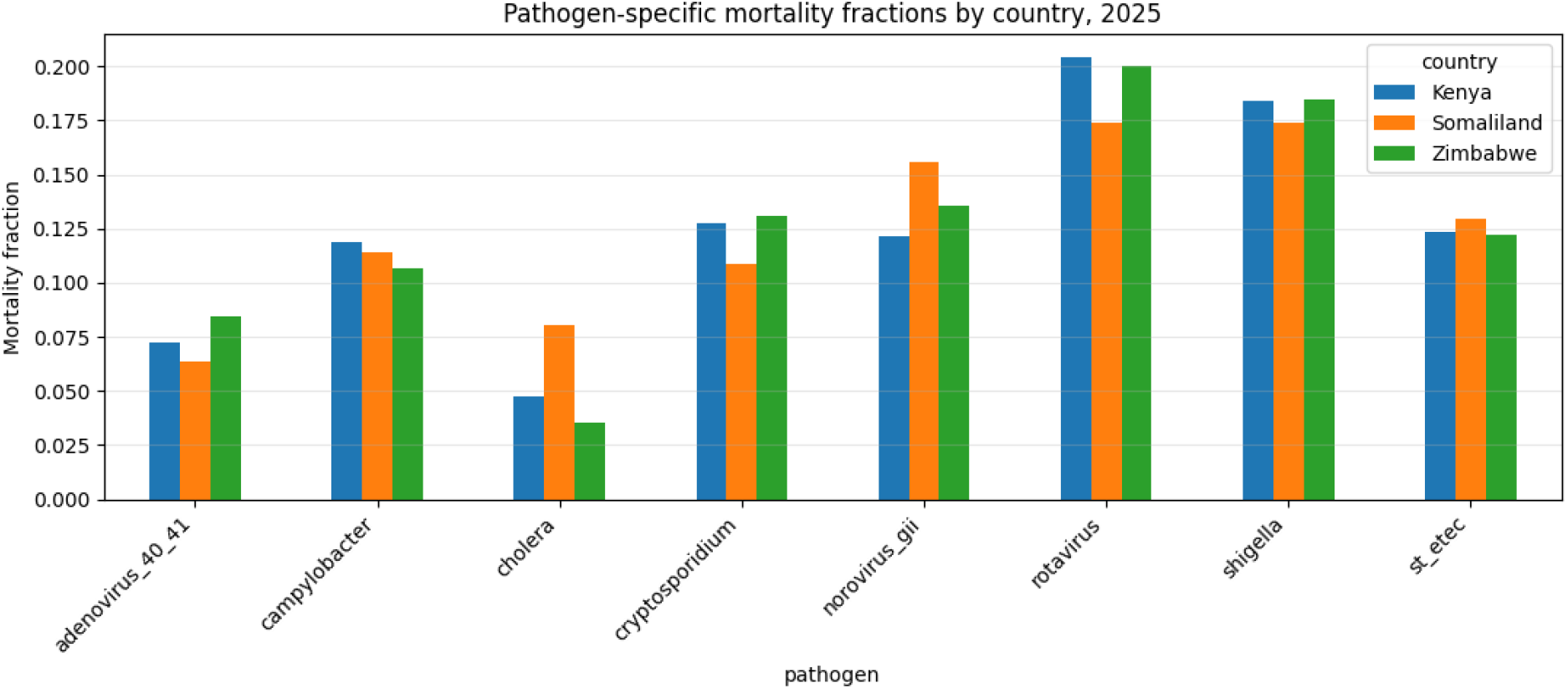
Proportion of under-five diarrheal deaths attributed to major enteric pathogens in Kenya, Somaliland, and Zimbabwe generated using the KESOZI Digital Twin framework.

### 3.11 Country-Specific Pathogen-Attributed Mortality Burden

Figure 12 presents the estimated pathogen-attributed diarrheal deaths among children under five across Kenya, Somaliland, and Zimbabwe in 2025. Rotavirus accounted for the highest mortality burden in all three countries, followed by Shigella. Cryptosporidium, Norovirus GII, and ST-ETEC contributed intermediate mortality burdens, whereas Cholera, Campylobacter, and Adenovirus 40/41 accounted for comparatively smaller numbers of attributed deaths. Kenya consistently exhibited the highest pathogen-attributed mortality across all pathogens, while Somaliland and Zimbabwe recorded lower but broadly similar mortality patterns.

**Figure 12.**
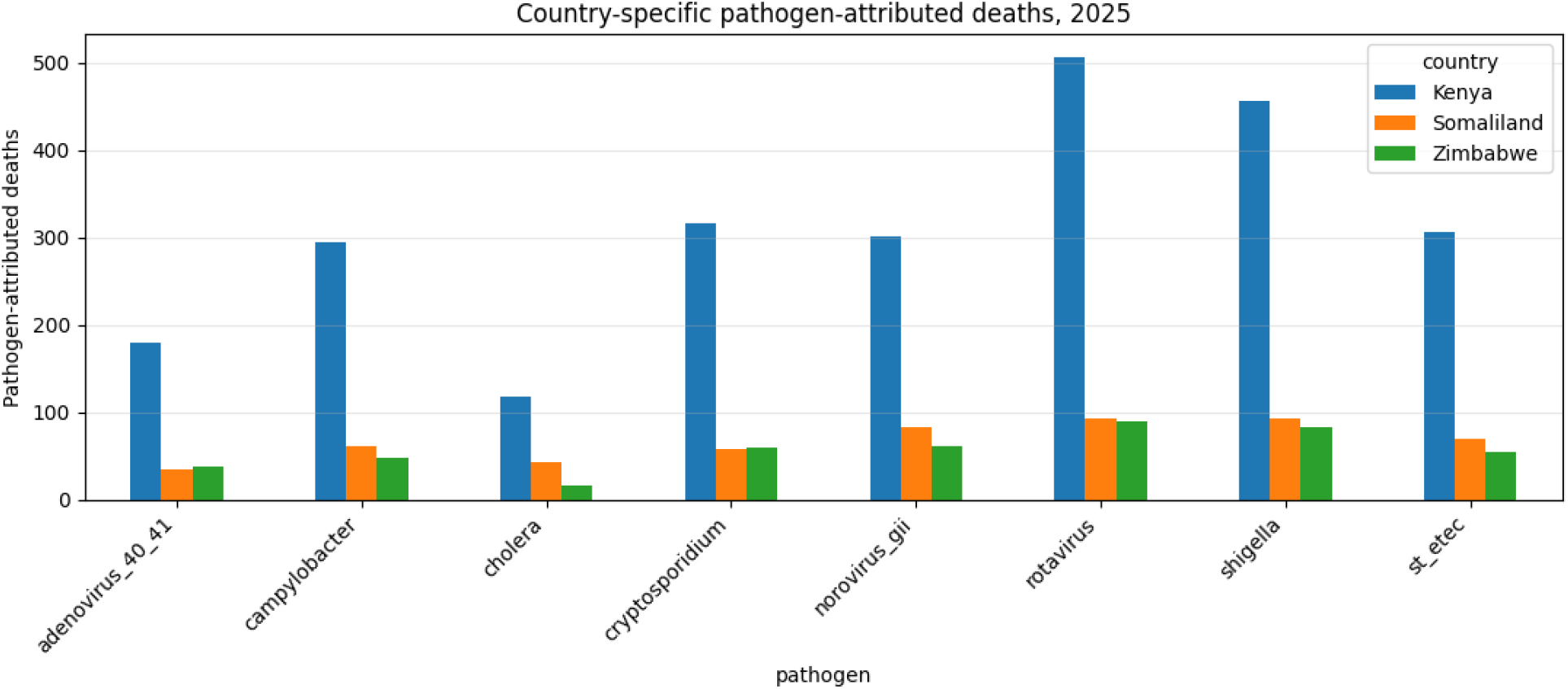
Estimated under-five diarrheal deaths attributed to major enteric pathogens in Kenya, Somaliland, and Zimbabwe generated using the KESOZI Digital Twin framework

### 3.12 Country-Level Mortality Uncertainty Assessment

Figure 13 presents the predicted under-five diarrheal mortality burden and corresponding 95% uncertainty intervals for Kenya, Somaliland, and Zimbabwe. Kenya exhibited the highest estimated mortality burden, with approximately 2,500 predicted deaths, followed by Somaliland and Zimbabwe with substantially lower mortality estimates. The uncertainty intervals were widest for Kenya, reflecting greater variability in the mortality estimates, while Somaliland and Zimbabwe showed narrower confidence intervals. Despite these differences, the uncertainty bounds remained relatively constrained, indicating stable model predictions across the study settings.

**Figure 13.**
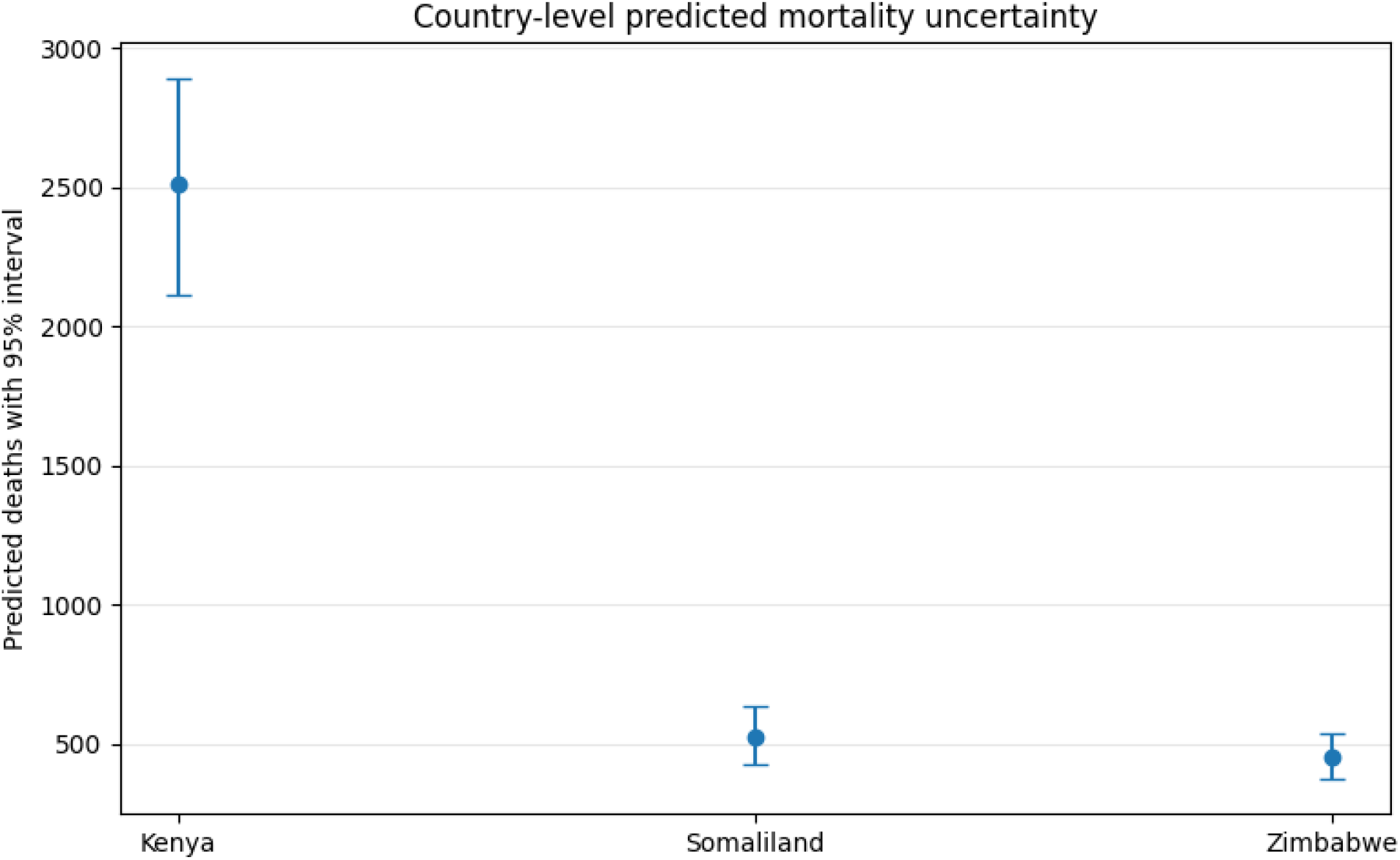
Country-level predicted diarrheal mortality with 95% uncertainty intervals in 2025. Estimated under-five diarrheal deaths and corresponding 95% uncertainty intervals for Kenya, Somaliland, and Zimbabwe generated using the KESOZI Digital Twin framework

### 3.13 Spatial Risk Network Analysis

Figure 14 presents a graph-based spatial risk feature derived from neighboring high-risk regions across Kenya, Somaliland, and Zimbabwe. The highest average synthetic mortality risk was observed in Las Anod (Somaliland), Matabeleland South (Zimbabwe), and Marsabit (Kenya), indicating that these locations were embedded within clusters of elevated disease burden. Other high-risk areas included Mandera and Samburu in Kenya and Erigavo in Somaliland. In contrast, Sheikh (Somaliland), Isiolo (Kenya), and Matabeleland North (Zimbabwe) exhibited comparatively lower neighborhood risk values. The spatial distribution demonstrates substantial heterogeneity in modeled mortality risk across regions and suggests the presence of geographically concentrated disease hotspots.

**Figure 14.**
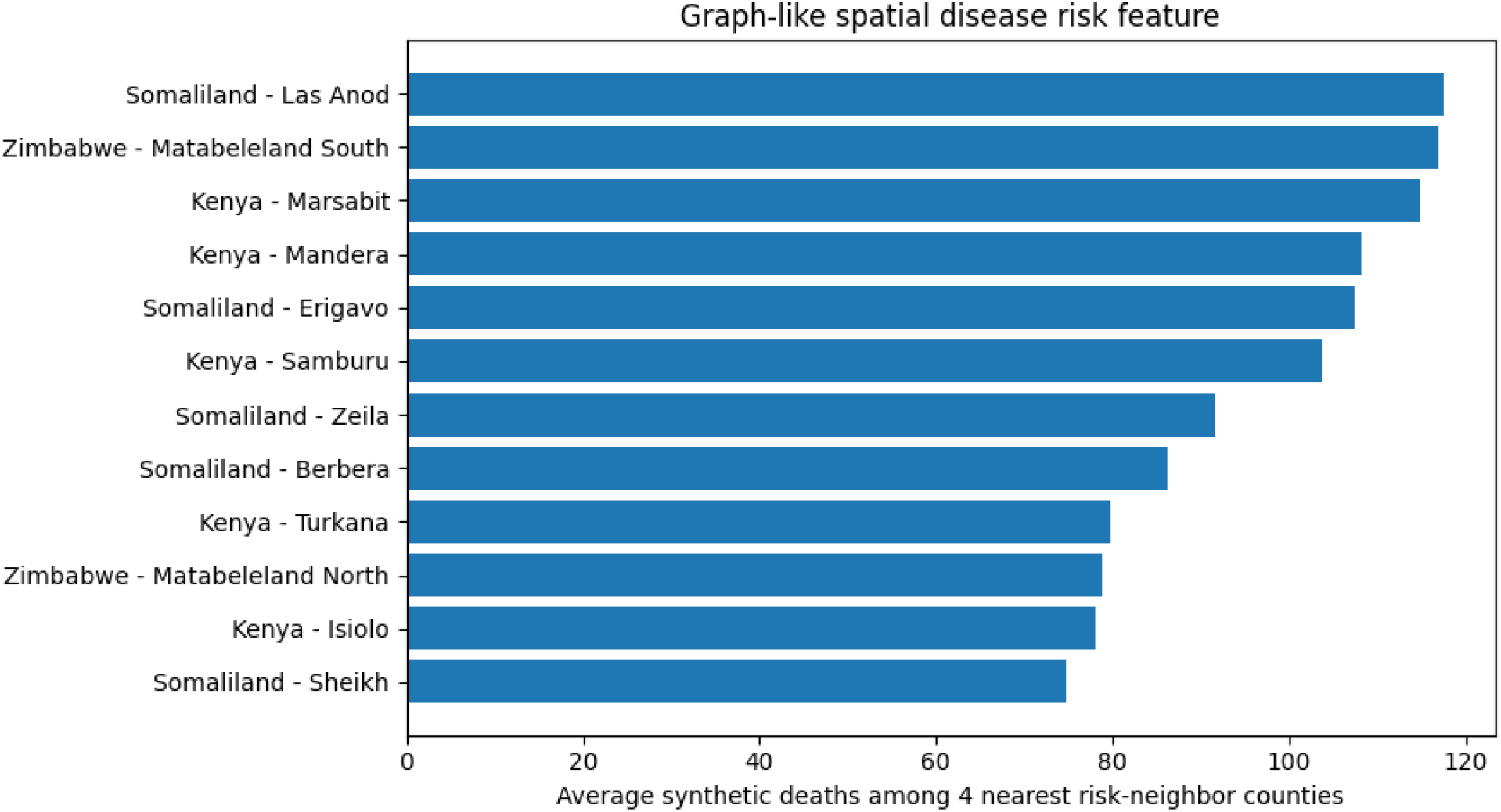
Graph-based spatial risk feature across study regions. Average synthetic diarrheal mortality risk among neighboring high-risk regions in Kenya, Somaliland, and Zimbabwe, illustrating geographic clustering of disease burden and potential transmission hotspots

### 3.14 Physics-Informed Neural Network (PINN) Trajectory Fitting

Figure 15 presents the Physics-Informed Neural Network (PINN) fit for the synthetic diarrheal infection trajectory in Kisumu, Kenya, from January 2024 to December 2025. The observed synthetic infection proxy exhibited substantial temporal variability, with several peaks and troughs reflecting fluctuations in disease transmission over time. In contrast, the PINN-generated trajectory produced a smooth underlying trend that captured the overall temporal dynamics while filtering short-term fluctuations and noise. The fitted trajectory remained relatively stable throughout the study period, suggesting that the PINN successfully learned the dominant temporal behavior of the synthetic disease process.

**Figure 15.**
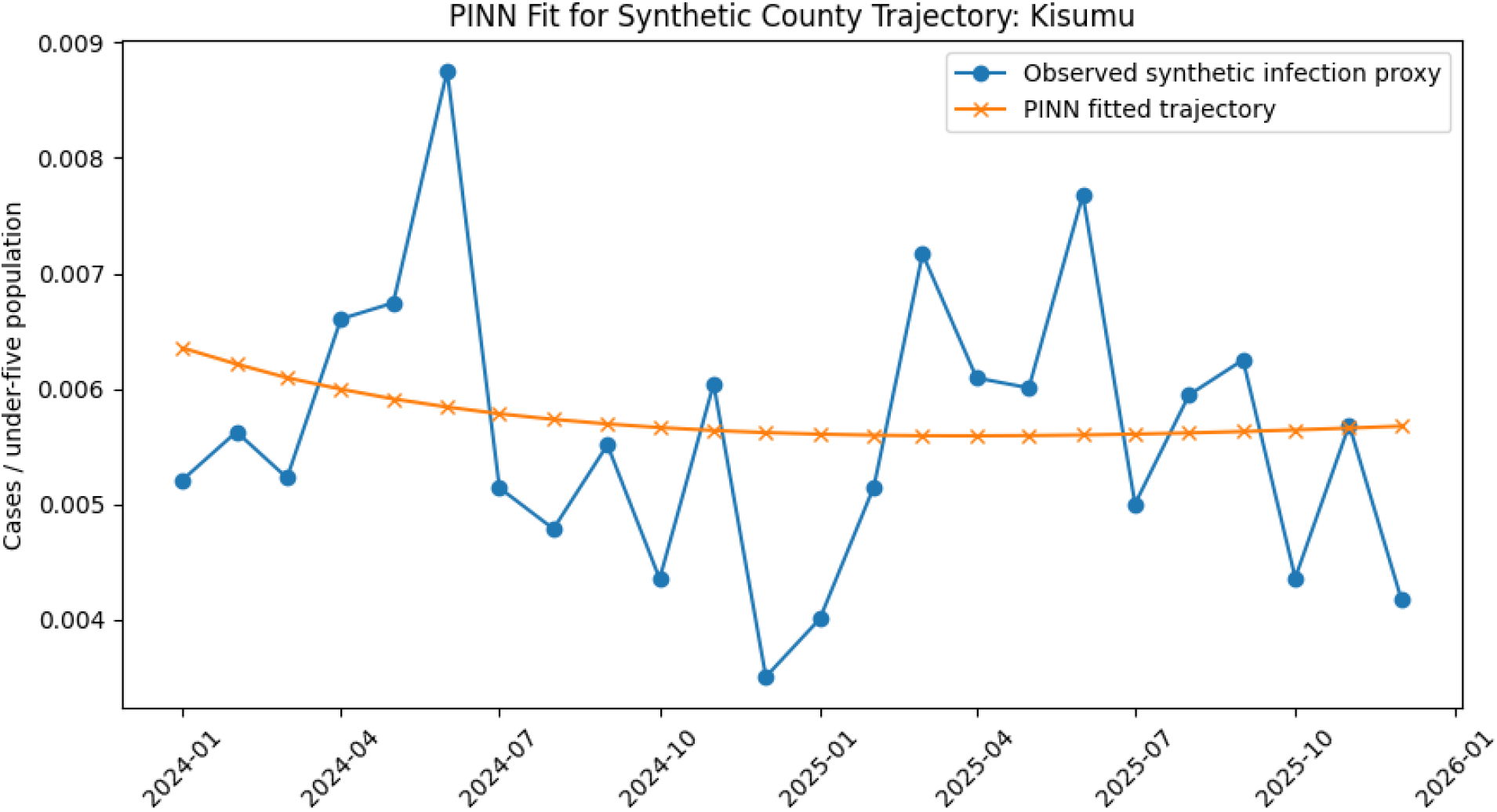
PINN fit for the synthetic diarrheal infection trajectory in Kisumu, Kenya (2024-2025). Observed synthetic infection proxy values and the corresponding Physics-Informed Neural Network fitted trajectory showing the underlying temporal trend in disease

### 3.15 PINN Training Convergence and Loss Dynamics

Figure 16 shows the evolution of the Physics-Informed Neural Network (PINN) training losses over 250 epochs. The total loss decreased rapidly during the initial training phase and continued to decline gradually throughout optimization, indicating stable convergence. The data loss exhibited the fastest reduction, falling by several orders of magnitude within the first 50 epochs before reaching a near-constant minimum value. Similarly, the physics loss decreased steadily over time, demonstrating improved adherence to the underlying governing constraints incorporated into the PINN framework. By the final training epoch, all loss components had stabilized at low values, indicating successful model training and convergence.

**Figure 16.**
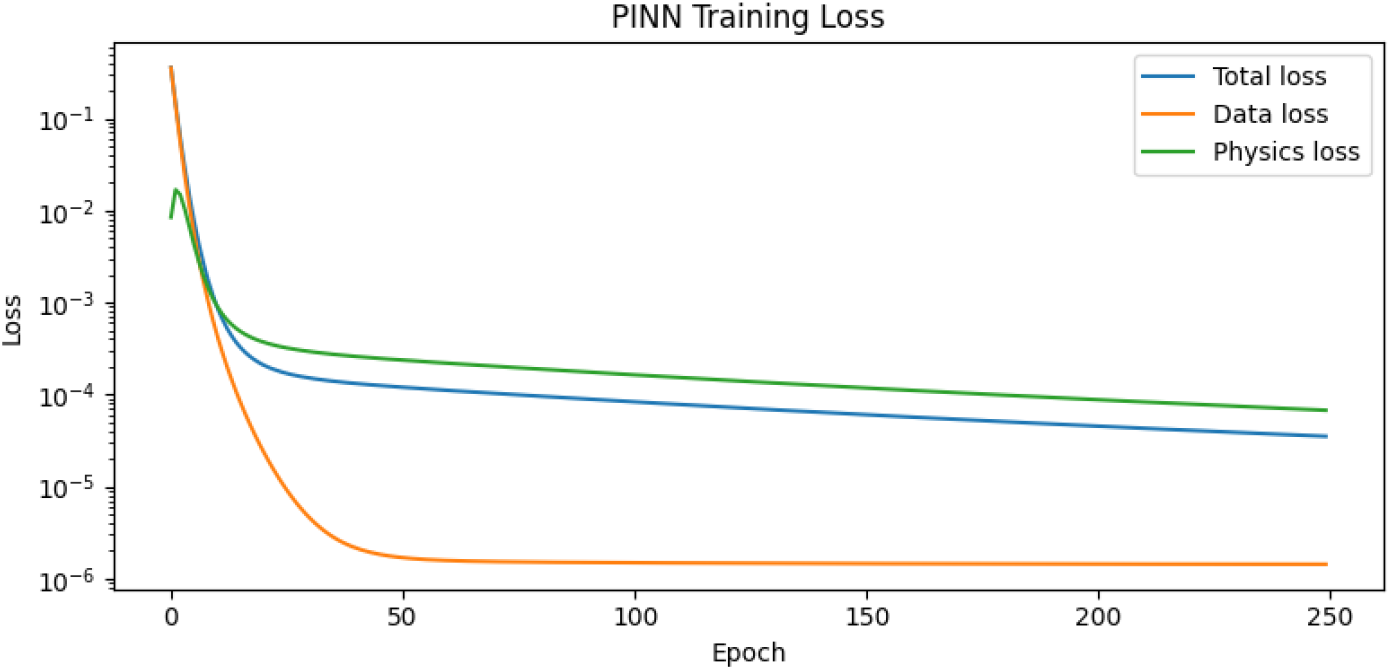
PINN training loss convergence over 250 epochs. Evolution of total loss, data loss, and physics loss during Physics-Informed Neural Network training, demonstrating stable convergence and improved adherence to both observational data and governing model constraints

### 3.16 Digital Twin Scenario Simulation

Figure 17 presents the county-level digital twin simulation for Kisumu under four intervention scenarios: baseline conditions, flood shock, WASH improvement, and combined WASH-plus-health intervention. The flood-shock scenario consistently produced the highest projected mortality burden throughout the 12-month simulation period, reflecting the adverse effects of environmental disruption on disease transmission. In contrast, the WASH-improvement scenario reduced projected deaths relative to baseline conditions, while the combined WASH-plus-health intervention achieved the greatest reduction in mortality across the simulation horizon. The separation between scenario trajectories became more pronounced over time, indicating cumulative benefits of preventive interventions.

**Figure 17.**
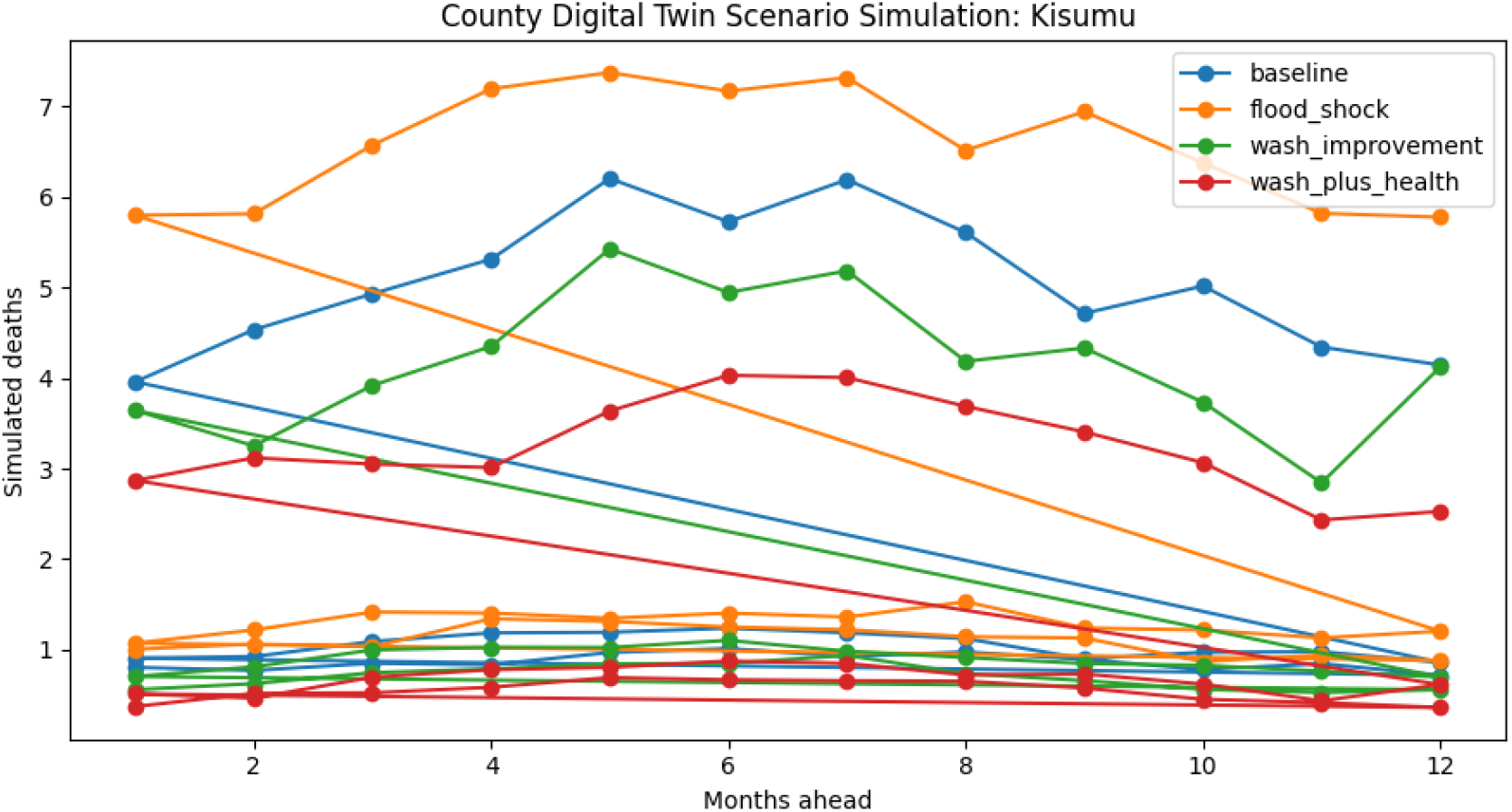
Digital twin simulation of intervention scenarios in Kisumu, Kenya. Projected under-five diarrheal mortality under baseline conditions, flood-shock conditions, WASH improvement, and combined WASH-plus-health interventions over a 12-month simulation period

### 3.17 Significance of the KESOZI Digital Twin Framework

The multimodal fusion framework improved diarrheal disease burden estimation relative to the Bayesian baseline by integrating temporal forecasting, machine-learning models, geospatial analytics, satellite-derived environmental intelligence, pathogen attribution, graph learning, and digital twin simulation. The framework successfully captured complex interactions among climate variability, unsafe-water exposure, sanitation vulnerability, healthcare access, pathogen prevalence, and nutritional risk. In addition, the uncertainty-calibration component generated stable prediction intervals, while the graph-learning framework identified spatial clustering patterns, transmission corridors, and potential outbreak hotspots across the study regions.

## 4. Discussion

The positive association between composite risk scores and diarrheal incidence demonstrates the ability of the KESOZI Digital Twin framework to integrate environmental, WASH, nutritional, and healthcare determinants into a meaningful measure of disease risk. The findings indicate that environmental vulnerability, inadequate sanitation, poor nutrition, and limited healthcare access collectively contribute to increased under-five diarrheal burden and support the use of risk-based approaches for identifying high-burden settings and prioritizing interventions. Climate forcing emerged as an important driver of disease burden, with temporal variability potentially reflecting changes in environmental contamination, pathogen survival, flooding, drought, and water-system disruptions. The differing climate patterns observed across Kenya, Somaliland, and Zimbabwe underscore the need for location-specific models that account for regional climatic dynamics when estimating diarrheal disease burden and planning interventions. Substantial heterogeneity was observed in mortality and morbidity burden across the study countries. Kenya exhibited the highest absolute mortality burden, whereas Somaliland showed the highest mortality and morbidity incidence rates, indicating greater population-level vulnerability. These findings demonstrate the importance of considering both absolute burden and population-adjusted indicators when assessing disease impact and allocating public health resources. The comparative burden profiles further highlight the capacity of the KESOZI Digital Twin framework to quantify multiple dimensions of disease burden simultaneously. Differences in risk-driver profiles revealed the multifactorial nature of diarrheal disease transmission. Somaliland’s elevated risk was associated with higher climate forcing, greater unsafe-water exposure, increased wasting prevalence, and reduced healthcare access, whereas Zimbabwe exhibited a comparatively lower vulnerability profile. Seasonal analyses further showed synchronized mid-year increases in diarrheal cases across the three countries, suggesting that environmental conditions strongly influence transmission dynamics and supporting the value of seasonal forecasting and early-warning systems.

Pathogen-attribution analyses consistently identified Rotavirus and Shigella as the dominant contributors to under-five diarrheal mortality, followed by Cryptosporidium and Norovirus GII. Similar pathogen rankings across countries suggest common transmission mechanisms, although differences in burden magnitude likely reflect variation in pathogen prevalence, healthcare access, environmental exposures, and nutritional status. These findings reinforce the importance of vaccination, WASH improvements, surveillance systems, and targeted clinical management for reducing pathogen-attributed mortality. The uncertainty analyses demonstrated that the KESOZI Digital Twin framework generated reliable mortality estimates while preserving country-level differences in disease burden. The wider uncertainty intervals observed for Kenya reflected greater variability in underlying risk factors, whereas Somaliland and Zimbabwe exhibited comparatively narrower prediction ranges. The inclusion of uncertainty quantification provides important information for evidence-based decision-making and resource allocation. Spatial risk network analysis highlighted the importance of geographic context in disease transmission. Regions with elevated neighborhood risk scores were characterized by overlapping environmental, WASH, nutritional, and healthcare vulnerabilities, while cross-border clustering patterns suggested that disease risk is influenced by shared ecological and transmission dynamics rather than administrative boundaries alone. These findings demonstrate the utility of graph-based approaches for hotspot detection, surveillance prioritization, and targeted intervention planning. The Physics-Informed Neural Network (PINN) successfully captured the underlying trajectory of the synthetic infection data while satisfying governing system constraints. Stable convergence of both data and physics losses indicated robust model training and demonstrated the potential of physics-informed learning for epidemiological forecasting, gap filling, and digital-twin applications under conditions of uncertainty and incomplete surveillance data.

Digital twin simulations further demonstrated the impact of alternative intervention scenarios on future mortality burden. Flood-related shocks substantially increased projected deaths, whereas improvements in WASH and healthcare access reduced mortality, with the combined intervention scenario producing the greatest health gains. These findings illustrate the value of digital twins for evaluating policy options, supporting proactive planning, and optimizing intervention strategies.

Collectively, the results demonstrate the value of integrating scientific machine learning, multimodal epidemiological modeling, graph learning, uncertainty quantification, and digital twin technologies within a unified surveillance framework. By capturing nonlinear epidemiological interactions, spatial transmission dynamics, and intervention effects, the KESOZI Digital Twin framework provides a scalable and policy-relevant platform for burden estimation, forecasting, outbreak preparedness, vaccine prioritization, WASH investment planning, climate-health adaptation, and evidence-based public health decision-making in data-constrained African settings.

## Author Approval Statement

All authors have read and approved the final version of this manuscript and consent to its submission to medRxiv. Each author has made a substantial contribution to the conception, design, data collection, analysis, interpretation, drafting, or revision of the work and agrees to be accountable for all aspects of the study. The authors confirm that the manuscript is original, has not been published elsewhere, and is not under consideration for publication by another journal or preprint server, except as permitted by medRxiv policies. All authors have reviewed the content and approved its public posting on medRxiv.

## Declaration

This study utilized publicly available, anonymized, synthetic, or secondary data sources and did not involve direct interaction with human participants or access to identifiable personal information. Therefore, ethics committee approval and informed consent were not required. Where applicable, all procedures were conducted in accordance with relevant institutional guidelines and regulations.

## Data Availability Statement

The datasets and materials used and/or analyzed during the current study are available from the corresponding author upon reasonable request. Publicly available data sources are appropriately cited within the manuscript.

## References

1. World Health Organization. (2024, March 7). Diarrhoeal disease. https://www.who.int/news-room/fact-sheets/detail/diarrhoeal-disease

2. Esteva, A., Robicquet, A., Ramsundar, B., Kuleshov, V., DePristo, M., Chou, K., Cui, C., Corrado, G., Thrun, S., & Dean, J. (2019). A guide to deep learning in healthcare. Nature Medicine, 25(1), 24–29. 10.1038/s41591-018-0316-z

3. Raissi, M., Perdikaris, P., & Karniadakis, G. E. (2019). Physics-informed neural networks: A deep learning framework for solving forward and inverse problems involving nonlinear partial differential equations. Journal of Computational Physics, 378, 686–707. 10.1016/j.jcp.2018.10.045

4. Kapoor, A., Ben, X., Liu, L., Perozzi, B., Barnes, M., Blais, M., & O’Banion, S. (2020). Examining COVID-19 forecasting using spatio-temporal graph neural networks. ICML Workshop on Graph Representation Learning and Beyond.

5. Fuller, A., Fan, Z., Day, C., & Barlow, C. (2020). Digital twin: Enabling technologies, challenges and open research. IEEE Access, 8, 108952–108971. 10.1109/ACCESS.2020.2998358

